# DNAm scores for serum GDF15 and NT-proBNP levels associate with a range of traits affecting the body and brain

**DOI:** 10.1101/2023.10.18.23297200

**Authors:** Danni A. Gadd, Hannah M. Smith, Donncha Mullin, Ola Chybowska, Robert F. Hillary, Dorien M Kimenai, Elena Bernabeu, Yipeng Cheng, Chloe Fawns-Ritchie, Archie Campbell, Danielle Page, Adele Taylor, Janie Corley, Maria Del C. Valdés-Hernández, Susana Muñoz Maniega, Mark E. Bastin, Joanna M. Wardlaw, Rosie M. Walker, Kathryn L. Evans, Andrew M. McIntosh, Caroline Hayward, Tom Russ, Sarah E. Harris, Paul Welsh, Naveed Sattar, Simon R. Cox, Daniel L. McCartney, Riccardo E. Marioni

**Author notes:** Corresponding author: Riccardo Marioni.

## Abstract

**Background:** Plasma growth differentiation factor 15 (GDF15) and N-terminal pro-B-type natriuretic peptide (NT-proBNP) are cardiovascular biomarkers that associate with a range of diseases. Epigenetic scores (EpiScores) for GDF15 and NT-proBNP may provide new routes for risk stratification.

**Results:** In the Generation Scotland cohort (N ≥ 16,963), GDF15 levels were associated with incident dementia, ischaemic stroke and type 2 diabetes, whereas NT-proBNP levels were associated with incident ischaemic heart disease, ischaemic stroke and type 2 diabetes (all P_FDR_ < 0.05). Bayesian Epigenome-wide association studies (EWAS) identified 12 and 4 DNA methylation (DNAm) CpG sites associated (Posterior Inclusion Probability [PIP] > 95%) with levels of GDF15 and NT-proBNP, respectively. EpiScores for GDF15 and NT-proBNP that were trained in a subset of the population. The GDF15 EpiScore replicated protein associations with incident dementia, type 2 diabetes and ischaemic stroke in the Generation Scotland test set (Hazard Ratios (HR) range 1.36 – 1.41, P_FDR_ <0.03). The EpiScore for NT-proBNP replicated the protein association with type 2 diabetes, but failed to replicate an association with ischaemic stroke. EpiScores explained comparable variance in protein levels across both the Generation Scotland test set and the external LBC1936 test cohort (R^2^ range of 5.7-12.2%). In LBC1936, both EpiScores were associated with indicators of poorer brain health. Neither EpiScore was associated with incident dementia in the LBC1936 population.

**Conclusions:** EpiScores for serum levels of GDF15 and Nt-proBNP associate with body and brain health traits. These EpiScores are provided as potential tools for disease risk stratification.

## Background

Delaying or preventing the onset of chronic diseases is a major challenge. Traditional risk factor models provide a foundation to achieve this (1,2), but can be augmented by molecular-level data. Plasma growth differentiation factor 15 (GDF15) and N-terminal pro-B-type natriuretic peptide (NT-proBNP) are biomarker candidates that have yielded promising results as indicators of a range of morbidities. GDF15 is associated with low-grade inflammation and age-related immunosuppression (3). Higher levels of GDF15 have been found, through Mendelian randomisation, to causally associate with increased risk of cardiometabolic stroke, atrial fibrillation, coronary artery disease and myocardial infarction (4). Two recent proteome-wide studies that assessed 1,301 (5) and 1,468 (6) proteins identified GDF15 as the top marker of multimorbidity. NT-proBNP is a metabolite of pro B-type natriuretic peptide (BNP), which is a natriuretic and diuretic hormone released by heart muscle in response to wall stretch (7). An inverse relationship between the levels of NT-proBNP in blood and incident diabetes has been reported (8), whereas lower levels of NT-proBNP have been associated with more favourable cardiovascular outcomes in randomised control trials (9–11). Elevated levels of GDF15 and NT-proBNP in individuals diagnosed with COVID-19 have been linked to more severe outcomes (12,13). Both protein markers have also been found to associate with vascular brain injury, poorer neurocognitive performance and incident dementias (14,15).

DNAm-based epigenetic scores (EpiScores) for blood proteins have been found to serve as markers of incident diseases (16) and augment clinically-used risk factors for risk stratification (17,18). DNAm reflects the body’s chronic response to low-grade inflammation, environmental and biological exposures (19–21). A study that directly compared an EpiScore for C-Reactive protein (CRP) to measured CRP found that the EpiScore had greater test-retest reliability over time point measures (22). This suggests that EpiScores may be more stable indictors than measured proteins in some instances. An EpiScore for GDF15 levels based on changes to DNAm at CpG sites across the genome is one of seven protein EpiScores that contribute to GrimAge, a leading epigenetic predictor of biological age acceleration, healthspan and lifespan (23,24). However, the performance of protein EpiScores against within-sample protein measurements in relation to incident diseases has not been comprehensively investigated. Additionally, EpiScores have typically been trained in samples of restricted size, with training sets typically ranging from 775 to 2,356 individuals (25) (23).

Here, we assess the viability of EpiScores for serum GDF15 and NT-proBNP as markers of disease outcomes and brain health (**Fig. 1**). Using GDF15 and NT-proBNP measures available in Generation Scotland (N ≥ 16,963), we first profile associations between GDF15 and NT-proBNP and four incident diseases (type 2 diabetes, ischaemic heart disease, ischaemic stroke and dementia), in addition to COVID-19 outcome severity. These diseases were chosen for the study as they have been linked to GDF15 and NT-proBNP and were available through electronic health linkage. We next map the epigenetic architectures of the two proteins, before training and testing protein EpiScores for them in subsets of Generation Scotland. In the test set, direct biomarker comparisons between measured proteins and the EpiScore equivalents are performed in relation to the four incident diseases assessed in the full Generation Scotland sample initially. EpiScores are then retrained in the full sample available and tested externally in the Lothian Birth Cohort 1936 (LBC1936), where associations with brain health traits are also profiled cross-sectionally and longitudinally.

**Figure 1.**
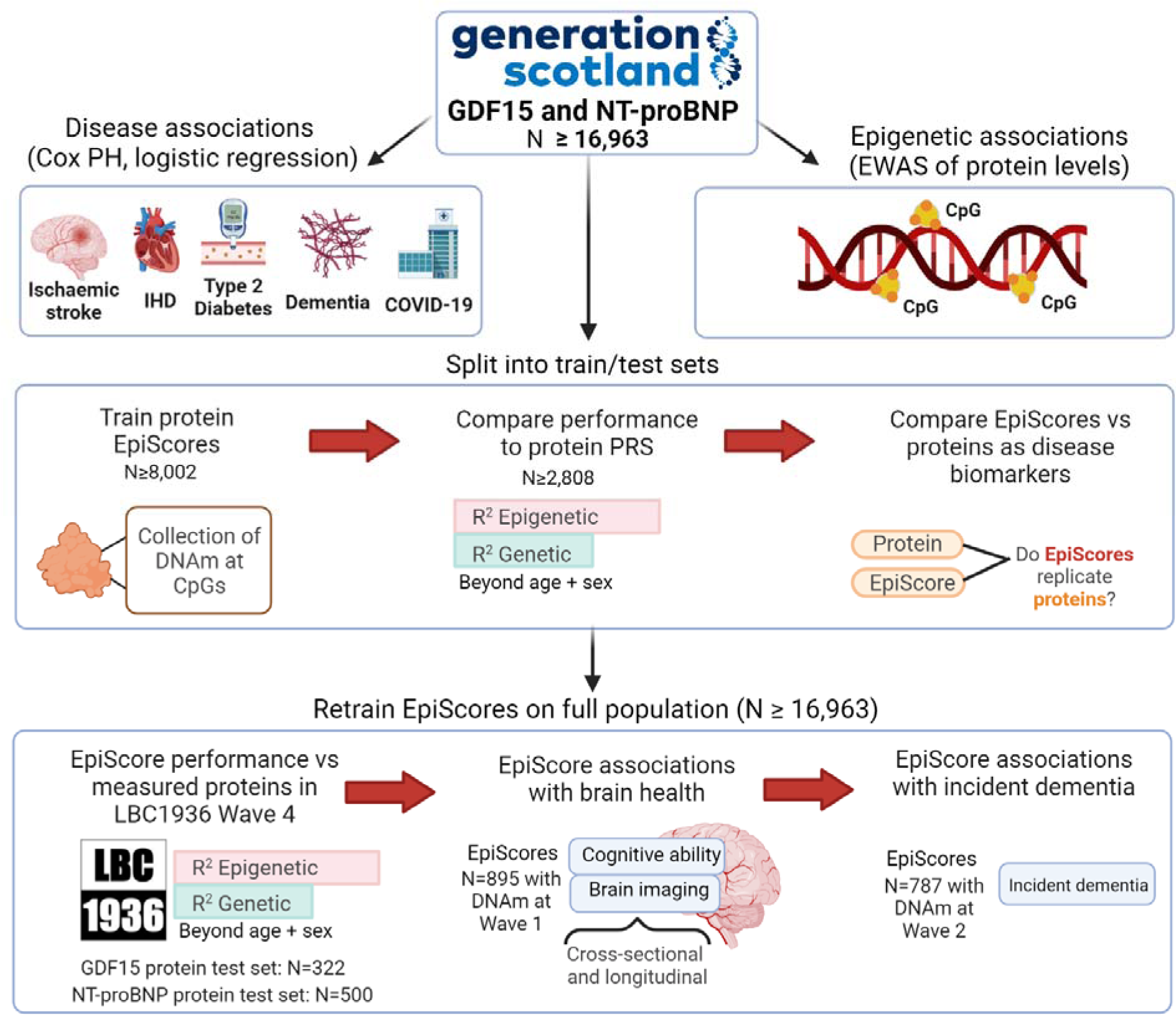
Study design for this assessment of GDF15 and NT-proBNP EpiScores as biomarkers. Disease associations and epigenome-wide association studies (EWAS) for each protein were first characterised in the full Generation Scotland sample. EpiScores for each protein were initially trained and tested in subsets of the population. This allowed EpiScores to be compared with measured proteins in associations with the four incident diseases profiled in the test set. EpiScores were then retrained on the full sample and tested externally in the LBC1936 Wave 4 population, which had measures of both proteins and DNAm available. EpiScores were projected into the larger LBC1936 Wave 1 population (that has DNAm but no protein measures) and profiled in associations with brain health traits, cross-sectionally and longitudinally. Consent for dementia linkage was available from Wave 2 of the LBC1936; therefore, we also tested whether EpiScores were associated with time-to-dementia. EpiScores were modelled with polygenic risk scores (PRS) for the proteins. CpG: cytosine-phosphate-guanine. IHD: ischaemic heart disease.

## Results

### Sample populations

There were 18,413 Generation Scotland participants (59% female) that had DNAm measurements, with a mean age of 48 years (SD 15), a minimum age of 17 years and maximum age of 98 years **(Supplementary Table 1)** (26,27). Of these, 17,489 had GDF15 measurements and 16,963 had NT-proBNP measurements. Subsets of this sample that were unrelated to one another were used to initially train and test EpiScores for GDF15 (N_train_ = 8,207, N_test_ = 2,954) and NT-proBNP (N_train_ = 8,002, N_test_ = 2,808) **(Supplementary Table 1)**. Measurements of serum GDF15 and NT-proBNP levels were available at Wave 4 (mean age 79 years, with 0.6 SD) of the LBC1936 study. These samples were used as external test sets for EpiScores trained on the full Generation Scotland sample. Of 507 individuals at Wave 4, 322 had GDF15 measures (48% female) and 500 had NT-proBNP measures (49% female). LBC1936 has successive Waves of measurements (Waves 1-5, collected at mean ages of 70, 73, 76, 79 and 82 years old, with SD < 1 at each Wave) (28,29). EpiScores were projected into Wave 1 (895 individuals with DNAm, but no protein measures) and evaluated in relation to cross-sectional and longitudinal brain health traits. As consent to dementia linkage was available from Wave 2, associations between EpiScores and time-to-dementia were also tested in LBC1936.

### GDF15 and NT-proBNP disease associations

Six associations (**Fig. 2**) were identified in Cox proportional hazards (PH) mixed effects models between protein levels and incident diseases in Generation Scotland (N ≥ 16,963). These associations had False Discovery Rate (FDR) P < 0.05 in basic (age and sex adjusted) models and P < 0.05 in fully-adjusted models (that further adjusted for smoking, alcohol intake, body mass index (BMI), social deprivation and years of education) **(Supplementary Table 2)**. Counts for cases, controls and mean time-to-onset for cases are provided in **Supplementary Table 2**. In basic logistic regression models, GDF15 was associated with subsequent hospitalisation due to COVID-19 (odds ratio (OR) per SD = 2.0, 95% confidence interval (CI) = [1.2, 3.4], FDR P = 0.037), as opposed to having COVID-19 without hospitalisation. An inverse association was identified between a one SD increase in NT-proBNP levels and COVID-19 hospitalisation (OR = 0.59, 95% confidence interval (CI) = [0.38, 0.93], FDR P = 0.046). No associations in relation to long-COVID as a binary outcome had FDR P < 0.05. Full summary statistics are provided in **Supplementary Table 3**.

**Figure 2.**
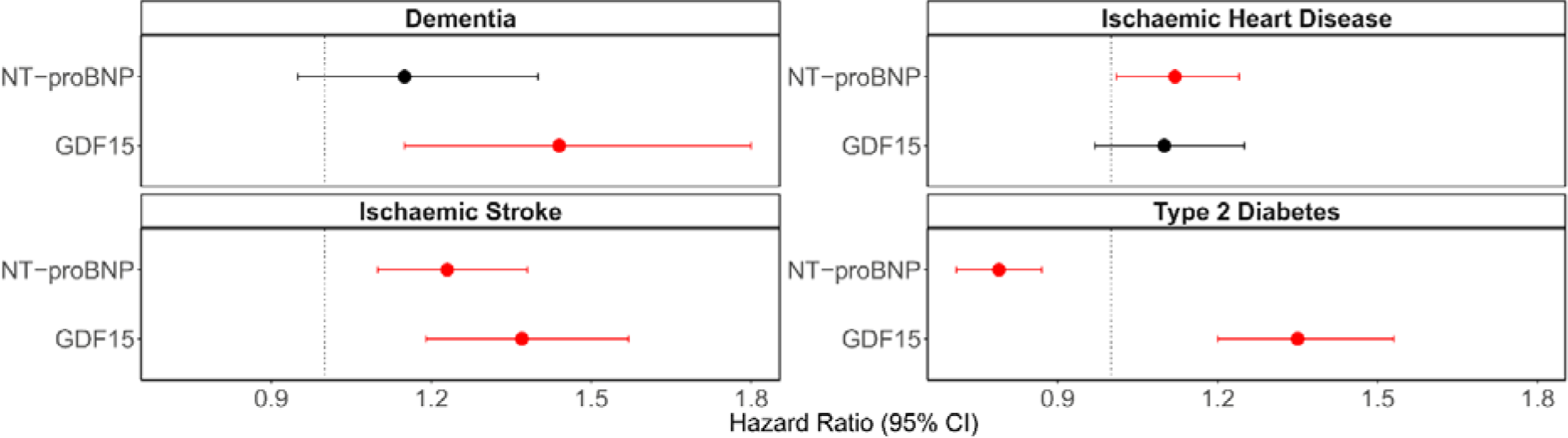
Disease associations for GDF15 and NT-proBNP in Generation Scotland (N ≥ 16,963). Fully-adjusted hazard ratios from Cox PH mixed effects regression models between protein levels and incident diseases are plotted with 95% confidence intervals. The six associations in red had FDR P < 0.05 in basic and P < 0.05 in fully-adjusted models, whereas associations that had P > 0.05 are shown in black. Hazard ratios are plotted per 1 SD increase in the rank-base inverse normalised levels of each marker. Fully-adjusted models controlled for age, sex, relatedness and common health and lifestyle factors (smoking, alcohol intake, BMI, social deprivation and years of education).

### GDF15 and NT-proBNP epigenetic associations

In variance components analysis of GDF15 and NT-proBNP in Generation Scotland (N ≥ 16,963), DNAm explained 36% of the variance in GDF15 levels (lower and upper confidence intervals [CIs] = 32% to 39%) and 32% of the variance in NT-proBNP levels (lower and upper CIs = 27% to 36%). In the EWAS, there were 12 and 4 associations (Bayesian Posterior Inclusion Probability [PIP] > 95%) between differential DNAm at 14 unique CpG sites and the levels of GDF15 and NT-proBNP, respectively. The CpG sites cg03546163 (*FKBP5*) and cg13108341 (*DNAH9*) were associated with both GDF15 and NT-proBNP. **Table 1** summarises the CpG sites, the biomarkers they associated with, the genes the CpGs are annotated to, and a selection of traits that DNAm at these CpGs have previously been associated with in EWAS studies. The full index of MRC IEU EWAS Catalogue associations (available as of August 2023) for these 14 CpG sites are available in **Supplementary Table 4**.

**Table 1.** EWAS of GDF15 and NT-proBNP levels in Generation Scotland (N ≥ 16,963). Posterior inclusion probabilities (PIPs) are provided for all CpG-protein associations (PIP > 0.95) in the BayesR EWAS. * Two CpGs were associated with both GDF15 and NT-proBNP. A selection of traits implicated in associations (P<3.8×10^−6^, with n>100) with the CpGs from the MRC-IEU EWAS Catalogue (as of August 2023) are shown. HbA1c: glycated haemoglobin. IgG: immunoglobulin G.

| CpG | PIP | Biomarker | CpG Gene | CpG trait associations (MRC-IEU EWAS Catalogue) |
| --- | --- | --- | --- | --- |
| cg03546163* | 0.98,<br>0.96 | GDF15,<br>NT-proBNP | *FKBP5* | Chronic kidney disease, fetal vs adult liver, body mass index, waist circumference, mortality, age, neurodegenerative disorders. |
| cg13108341* | 1.00,<br>0.98 | GDF15,<br>NT-proBNP | *DNAH9* | Cancer treatment |
| cg00757033 | 1.00 | NT-proBNP | *WDR51B* | Crohn's disease, inflammatory bowel disease, age |
| cg05412028 | 0.99 | NT-proBNP | *ABCC4* | Age, ageing, primary Sjogrens syndrome |
| cg19693031 | 1.00 | GDF15 | *TXNIP* | Fetal vs adult liver, triglycerides, sex, hbA1c, alcohol consumption, blood pressures, hepatic fat, waist circumference, cholesterol measures, age. |
| cg02650017 | 1.00 | GDF15 | *PHOSPHO1* | Type 2 diabetes, primary Sjogrens syndrome, C-reactive protein, body mass index, serum high-density lipoprotein cholesterol, Crohn's disease, body mass index, coagulation factor VIII, eosinophilia, age. |
| cg06918740 | 1.00 | GDF15 | N/A | N/A |
| cg08900409 | 1.00 | GDF15 | *PGPEP1* | Age |
| cg25460262 | 1.00 | GDF15 | *GDF15* | Fetal vs adult liver |
| cg21088460 | 1.00 | GDF15 | *GDF15* | N/A |
| cg05575921 | 1.00 | GDF15 | *AHRR* | Extensive set of smoking-associated traits, lung function/cancer traits, body mass index, serum cotinine, C-reactive protein, IgG glycosylation measures, educational attainment, cognitive ability, statin use, urinary cadmium, mortality, post-traumatic stress disorder, age, acute myocardial infarction. |
| cg25410121 | 1.00 | GDF15 | N/A | N/A |
| cg15058033 | 0.97 | GDF15 | *PLXNB2* | N/A |
| cg16993186 | 0.97 | GDF15 | *CELF2* | N/A |

### EpiScores for GDF15 and NT-proBNP within Generation Scotland

EpiScores for GDF15 and NT-proBNP were initially trained and tested in subsets of Generation Scotland that were unrelated to one another. Predictor weights for EpiScores are available in **Supplementary Table 5**. Performance in the test set was modelled through the incremental variance (R^2^) in protein levels that scores could explain beyond a null linear regression model that adjusted for age and sex. The EpiScore for GDF15 trained using the full set of EPIC array probes had an R^2^ of 12.2%, whereas the NT-proBNP EpiScore had an R^2^ of 5.7%. Similar performance was observed when comparing with EpiScores trained using sites available on the 450k array subset **(Supplementary Fig. 1**). When modelling EpiScores and polygenic risk scores (PRS) derived for each protein **(see Methods),** additive effects beyond the null model were observed for GDF15 (R^2^ of 15.5%) and NT-proBNP (R^2^ of 6.9%). A full summary of the results is provided in **Supplementary Table 6.**

### EpiScore replication of protein biomarker associations within Generation Scotland

The same Cox PH model structure (as shown in **Fig. 2**) was used to directly compare protein levels and EpiScores in the Generation Scotland test. All protein-disease associations – except the association between NT-proBNP and ischaemic stroke – were replicated by EpiScores in fully-adjusted models (**Fig. 3, Supplementary Table 7)**. Mean time-to-onset, counts for cases and controls and full summary statistics are available in **Supplementary Table 7.** Mean attenuation in the absolute log of the HR due to the additional adjustment for lifestyle factors beyond age and sex was 6% for protein associations and 12% for EpiScore associations. Of the four protein EpiScore associations identified in fully-adjusted models, three withstood further adjustment for estimated immune cell proportions (attenuation in the absolute log of the HR ranging from 0 – 9%). The association between the GDF15 EpiScore and dementia had P = 0.064, with 5% attenuation in the absolute log of the HR. **Supplementary Table 7)**. As only five instances of COVID-19 hospitalisation and nine instances of long-COVID were reported in the test population, we did not conduct protein EpiScore and protein comparisons for these outcomes.

**Figure 3.**
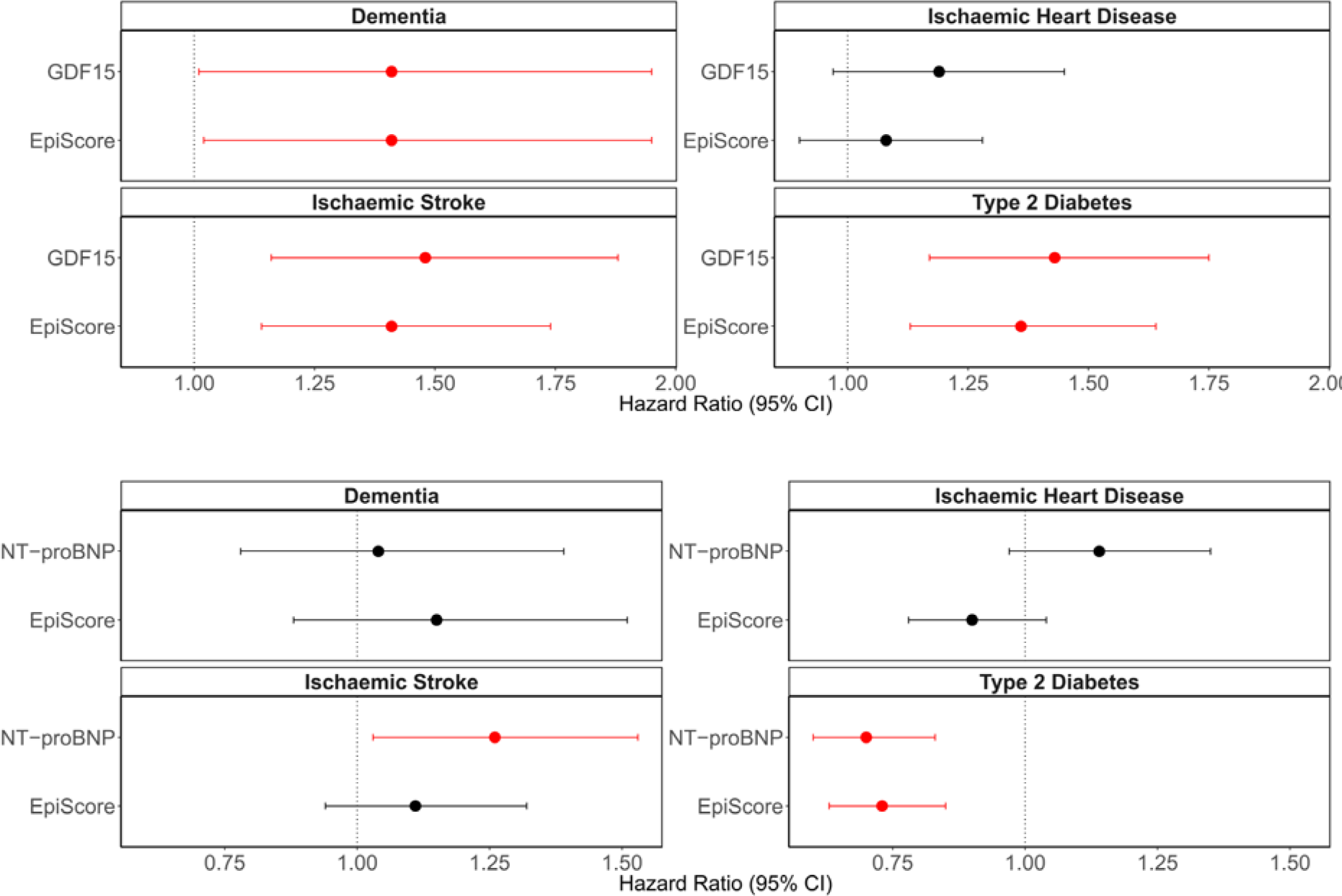
Comparison of EpiScores versus measured protein equivalents in fully-adjusted associations with incident diseases in the Generation Scotland test sample (N ≥ 2,808). For each disease, the protein-disease association is plotted, with the equivalent protein EpiScore-disease association shown directly beneath it for comparison. Hazard ratios are plotted per 1 SD increase in the rank-based inverse normalised levels of each marker. Nine associations (red) had FDR P < 0.05 in basic and P < 0.05 in fully-adjusted Cox proportional hazards mixed effects models in the test samples. Fully-adjusted models adjusted for age, sex, relatedness and common lifestyle risk factors (smoking, alcohol intake, BMI, social deprivation and years of education). Associations that were non-significant (P > 0.05 in fully-adjusted models) are shown in black.

### EpiScore application to the LBC1936 external cohort

EpiScores for each protein were then retrained in the entire Generation Scotland sample (N_GDF15_ = 17,489 and N_Nt-proBNP_ = 16,963). Although we make predictor weights for EpiScores trained on the full EPIC array probes and the subset of probes on the older 450k array available (**Supplementary Table 8**), the LBC1936 external test cohort in this study measured DNAm using the 450k array. Thus, the EpiScores trained on the 450k probe subset were projected into this population for external validation.

In the LBC1936 test sample (N_GDF15_ = 322 and N_Nt-proBNP_ = 500), incremental R^2^ values of 8.9% for GDF15 and 8.1% for NT-proBNP EpiScores were observed, beyond age and sex-adjusted linear regression models (**Fig. 4a**). When a PRS for each protein was modelled together with the EpiScores, incremental variance explained rose to 13.7% and 9.1% for GDF15 and NT-proBNP, respectively **(Supplementary Table 9)**. Finally, the GDF15 EpiScore generated previously by Lu *et al* as part of the GrimAge biological age acceleration predictor (23) was projected into the Wave 4 GDF15 test set (322 individuals) and evaluated. It explained 5.6% of the variance in GDF15 beyond age and sex, as compared to the 8.9% observed modelling our updated GDF15 score. The two GDF15 EpiScores had a Pearson correlation *r* = 0.32 in the test sample.

**Figure 4.**
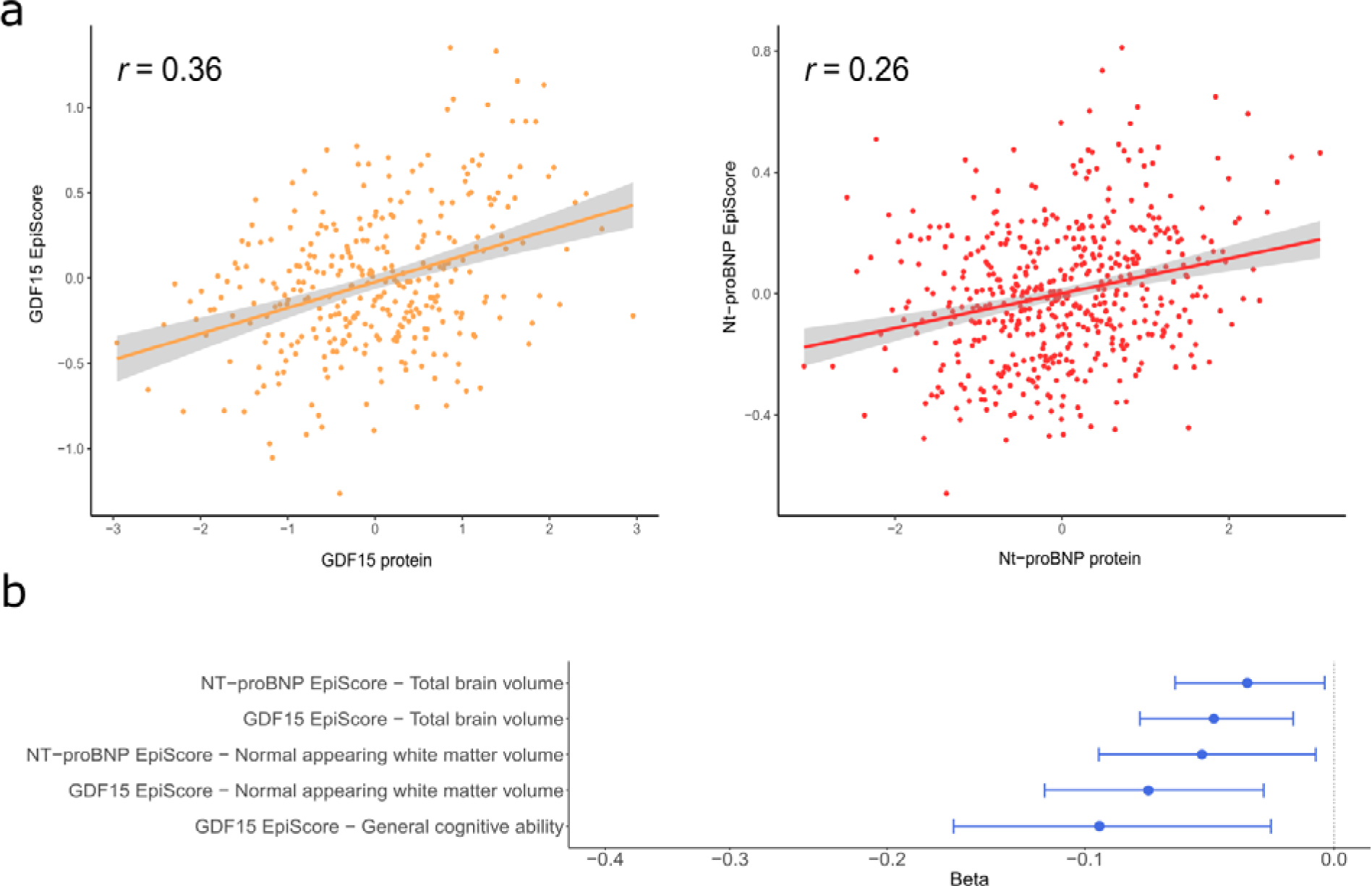
External assessment of the GDF15 and NT-proBNP EpiScores in LBC1936. **a,** Correlation plots between measured protein levels and GDF15 (orange) and NT-proBNP (red) EpiScores in the LBC1936 Wave 4 external test set (N_GDF_ = 322, N_NT-proBNP_ = 500). Pearson correlation coefficients are annotated in each instance. **b,** Standardised beta coefficients derived from structural equation models (SEMs) between EpiScore levels at LBC1936 Wave 1 (N=895 with DNAm, N=1,091 total) and cross-sectional measures of brain health traits that had FDR P < 0.05 in basic (age and sex adjusted) models and P < 0.05 after adjustment for further lifestyle covariates. All associations had a negative beta coefficient (blue). Twenty EpiScore-trait associations were tested in total: 10 cross-sectionally and 10 assessing longitudinal change in brain traits.

### EpiScore associations with brain health traits in LBC1936

The EpiScores that were validated against protein measures in the LBC1936 Wave 4 external test set were then projected into methylation measured at Wave 1 (a time point nine years prior), which has a larger DNAm sample available but no protein measures. Structural equation models were then run to characterise associations between the protein EpiScores and five brain health traits (cognitive ability and four structural brain imaging measures). This allowed for EpiScore relationships with both cross-sectional brain health (Wave 1, N=895 individuals with EpiScores, total model N=1,091) and longitudinal change in brain health (Waves 1-5 for cognitive change and Waves 2-5 for brain imaging changes) to be tested (five brain health traits x two EpiScores x cross-sectional and longitudinal associations = 20 hypothesis tests).

Seven of the twenty basic model associations tested had FDR P < 0.05 **(Supplementary Table 10)**. All seven associations involved cross-sectional brain health phenotypes and had negative effect estimates (standardised betas ranging from −0.05 to −0.19). Higher GDF15 and NT-proBNP EpiScores were associated with lower general cognitive ability and lower brain volumes. None of the ten slope associations assessing relationships between the EpiScores and longitudinal change in the five brain health phenotypes were significant at FDR P < 0.05. In models that further adjusted for additional health and lifestyle factors, five associations had P < 0.05 **(Fig.4b, Supplementary Table 10)**. A one standard deviation increase in GDF15 EpiScore levels was associated with lower normal appearing white matter volume (Beta = −0.07, SE = 0.02, P = 2.2×10^−3^), poorer general cognitive ability (Beta = −0.09, SE = 0.04, P = 9.1×10^−3^) and lower total brain volume (Beta = −0.05, SE = 0.02, P = 3.5×10^−3^). A one standard deviation increase in NT-proBNP EpiScore levels was associated with lower normal appearing white matter volume (Beta = −0.05, SE = 0.02, P = 0.02) and lower total brain volume (Beta = −0.03, SE = 0.01, P = 0.03). In a sensitivity analysis that further adjusted for immune cell proportions, the two NT-proBNP associations were attenuated (P > 0.07) and the associations between NT-proBNP and lower cognitive ability was found to be significant (Beta = −0.10, SE = 0.04, P = 6.0×10^−3^). In the sensitivity analysis, the three GDF15 associations remained significant (P < 0.05), with a mean attenuation of 11% in Beta effect magnitude **(Supplementary Table 10)**.

Individuals consented to share disease information from electronic health records from Wave 2 of the study onwards. In Cox regression models that utilised Wave 2 as the baseline and modelled incident dementia as the outcome (N_cases_ = 108, N_controls_ = 672, mean time-to-event for cases = 8.6 years [SD 3.42] and maximum follow-up of 14.3 years), no associations were identified for either EpiScore **(Supplementary Table 11).**

## Discussion

Here, biomarker-disease associations for GDF15 and NT-proBNP were first observed in Generation Scotland, prior to developing EpiScores for these proteins. EpiScores replicated protein associations with incident diseases in the Generation Scotland test sample. In the LBC1936 external test population, the GDF15 and NT-proBNP EpiScores explained 9% and 8% of the variance in the protein levels, respectively, with higher levels of the EpiScores associated with poorer brain health cross-sectionally. EWAS of each protein highlighted 14 CpGs with differential DNAm.

This study provides EpiScores for GDF15 and NT-proBNP trained in the largest samples to date as tools for health stratification. Despite the LBC1936 test set being older than the Generation Scotland cohort (mean age of 79 *versus* 48 years), the EpiScores had R^2^ values comparable to those observed in the Generation Scotland test set. In the external LBC1936 test set, the GDF15 EpiScore had improved performance (an additional R^2^ of 3.3%) when compared to the GDF15 EpiScore derived by Lu *et al* in 2019 as part of the GrimAge biological age acceleration predictor (23). This is likely due to differences in the sample sizes used for training the two GDF15 scores (2,356 individuals as compared to 17,489 individuals in our study). It may also be due to our training and testing populations having more homogeneous ancestry (Scottish) than the populations used to train the original GrimAge GDF15 score (mixed white European ancestry). No other EpiScores for either GDF15 or NT-proBNP exist in the literature to our knowledge; these EpiScores can therefore be utilised as new tools for risk stratification and can be projected into any cohort with Illumina-based DNAm. We provide EpiScore weights trained on both the 450k and EPIC arrays for use in future research.

Generation Scotland is one of the world’s largest single cohort resources with DNAm, protein measures, and extant data linkage to electronic health records. This allowed for direct comparisons between protein and EpiScore measures in the context of incident disease analyses, which has only recently been possible owing to the expansion of the cohort’s epigenetic resource. As DNAm may record chronic exposure to a range of environmental risk factors (30) and biological processes such as inflammation (20,31), EpiScores may be reflective of a range of biological pathways that occur upstream of disease diagnoses. Given that GDF15 and Nt-pro-BNP are promising biomarkers for a range of diseases, our EpiScores are well-positioned candidates with many potential use-cases. The results of inclusion of the PRS for proteins in incremental variance models suggested that EpiScore signals were largely independent of genetic architectures on the proteins, as additive improvements in incremental variance observed when PRS and EpiScores were modelled together. This is in concordance with previous studies that found additive epi/genetic heritability estimates for plasma protein levels (32,33). While we have previously regressed out protein quantitative trait loci (pQTLs) from proteins prior to training EpiScores (25), there is an argument that EpiScores capturing a combination of genetic and epigenetic signatures may enhance the disease-predictive signal available. Both approaches are likely viable for the creation of new biomarkers.

The higher proportion of variance explained by the GDF15 EpiScore as compared to the NT-proBNP EpiScore suggests that GDF15 was better-characterised by DNAm differences across the genome. This may be due to its association with chronic inflammation, as we have observed particularly strong DNAm signatures associated with inflammatory proteins in previous work (25,34). A stronger DNAm signature was also observed for GDF15 in our EWAS analyses. To our knowledge, this represents the first EWAS of NT-proBNP. The only other EWAS of GDF15 levels was performed by us previously, using GDF15 measures from the SomaLogic assay (34), where we identified no associations for GDF15 passing Bonferroni correction. The improved power to detect associations in the present study (17,489 rather than 774 individuals) may have facilitated identification of associations in the present study. There were two CpG sites associated with both GDF15 and NT-proBNP (cg03546163 in *FKBP5* and cg13108341 in *DNAH9*), which suggests a partially-shared DNAm signature across the proteins. FK506-binding protein 5 (FKBP5) is implicated in cellular stress response (35). One previous study found cg03546163 to be differentially methylated in 107 individuals with type 2 diabetes that went onto develop end stage renal disease, versus 253 controls who did not (36).

The lack of associations with incident ischaemic heart disease in the Generation Scotland test set may be due to limited sample size, as an association between protein NT-proBNP and ischaemic heart disease was observed in the full Generation Scotland sample. Additionally, the GDF15 EpiScore association with incident dementia observed in Generation Scotland did not replicate in LBC1936. This may be due to differences in the way the phenotypes were defined across LBC1936 (consensus committee) versus Generation Scotland (Read and ICD codes only), or different cohort sampling frames and recruitment strategies.

Our findings support previous work identifying associations between GDF15 and Nt-proBNP protein levels and severe COVID-19 outcomes in hospitalised individuals (12,13). Although very few hospitalisation cases were available (n=28), both proteins (sampled a mean of 11 years prior to COVID-19 diagnoses) associated with subsequent hospitalisation due to COVID-19. GDF15 is likely to be elevated in individuals with multiple morbidities that may contribute towards greater risk of hospitalisation due to viral illnesses. Diabetes has been associated with increased risk of hospitalisation and adverse outcomes in COVID-19 (37,38). Both proteins (and equivalent EpiScores) should be investigated in populations that have DNAm quantified nearer to, or at COVID-19 diagnoses to further resolve these signals.

This study has several limitations. First, whereas it was advantageous to have common ancestry across both (Scottish) training and validation datasets, future studies should test EpiScores across larger populations that include non-European ancestries and larger age ranges as data become available. Second, emerging evidence has quantified differences in genetic associations with the measurements of the same proteins across panels that use antibody-based versus aptamer-based quantification methods (39). A particular example of interest from this study was GDF15 levels, which was highlighted as a protein that may have different conformational shapes (isoforms) that are targeted by the two assay methods (39). While it is likely that increased training sample size led to improved performance of our GDF15 score versus the GrimAge GDF15 score in the LBC1936 test set, it is possible that technical or biological variability across protein assays may also underlie differences in performance of scores. EpiScores trained on protein measurements from different panels should be therefore be compared further when data become available. Similarly, differences in the protein assay method across the previous EWAS of GDF15 (aptamer-based) that we ran and the present study (immunochemiluminescence) may also introduce variability and EWAS across multiple protein panels should be compared when samples are available.

## Conclusions

In conclusion, EpiScores for blood-based GDF15 and NT-proBNP levels are generated in this study and have been found to be useful indictors of disease risk stratification, with disease-specific use-cases. The EpiScores can be derived in any population with Illumina-based DNAm measurements and may be integrated into epigenetic screening panels in future to better-identify high-risk individuals.

## Methods

### Generation Scotland

Generation Scotland is a population-based cohort study that includes ∼8,000 families from across Scotland (26,27). Study recruitment of 23,960 participants occurred between 2006 and 2011, while participants were aged between 18 and 99 years. In addition to completing health and lifestyle questionnaires, participants donated blood samples for biomarker and omics measurement. Details on DNAm quality control in Generation Scotland are provided in **Supplementary Information**. The quality-controlled DNAm dataset comprised a total of 18,413 individuals with 760,838 CpG sites available on the EPIC array. GDF15 and NT-proBNP measurement details are provided in **Supplementary Information.** There were 18,413 individuals with GDF15 measures **(Supplementary Table 1)** and these were subset to 17,489 individuals that had DNAm (mean 1038.7 pg/mL [SD 928]). There were 17,863 individuals with NT-proBNP measures, with 16,963 that had DNAm available (mean 94.6 pg/mL [SD 211.2]). Electronic health records via data linkage to GP records (Read 2 codes) and hospital records (10^th^ revision of the International Classification of Diseases codes [ICD-10 codes]) were assessed prospectively from the time of blood draw. Incident data for all-cause dementia, type 2 diabetes, ischaemic stroke and ischaemic heart disease were considered with censoring date October 2020. Dementia cases were defined as per a previous review of dementia linkage codes (40), whereas code lists for all other diseases are available in **Supplementary Tables 12-14**. Prevalent cases (ascertained via retrospective linkage or self-report from a baseline questionnaire) were excluded from each disease trait, leaving only incident diagnoses. Dementia analyses were limited to cases/controls with age of event/censoring ≥ 65 years. Type 1 and juvenile diabetes cases were treated as control observations in the type 2 diabetes analyses. Death was treated as a censoring event.

### Lothian Birth Cohort 1936

The Lothian Birth Cohort 1936 (LBC1936) is a longitudinal study of ageing of people residing in Edinburgh and surrounding areas in Scotland (N = 1,091) (28,29). Individuals were born in 1936 and completed an intelligence test when they were 11 years old. They were later recruited to the cohort at a mean age of 70 years old and have been followed up triennially for a series of cognitive, clinical, social and physical measurements in five Waves (mean ages 70, 73, 76, 79 and 82 – all with SD below 1 for measures at each Wave). Blood samples were taken and used to derive protein, epigenetic and genetic measurements. Sample measurement details for the DNAm measures available in LBC1936 are provided in **Supplementary Information**. DNAm is available at the four successive waves of the study (N=895, 787, 619 and 507 in Waves 1, 2, 3 and 4, respectively). Both GDF15 (N=322) and NT-proBNP (N=500) serum levels were measured at Wave 4 of the study (mean age 79 years, SD 0.6) and were used to externally test EpiScore performance. From Wave 2 of the LBC1936, individuals consented for linkage to health records for research. Dementia cases were defined by a consensus committee that completed decisions in August 2022 (41). Potential cases were identified through a combination of electronic health record linkage, death certificate data and clinician visits to individuals that were suspected of having cognitive impairments or dementia. Of the 865 individuals who had provided linkage consent at Wave 2, 118 were confirmed as having dementia.

### Epigenome-wide association studies in Generation Scotland

The xBayesR+ software implements penalised Bayesian regression on complex traits (30). The BayesR method has been found to outperform linear and mixed model approaches and implicitly adjusts for probe correlations, data structure (such as relatedness) and unobserved confounders (30,32). Prior mixture variances for the methylation data (760,838 CpG sites) were set to 0.001, 0.01 and 0.1 and epigenome-wide associations studies (EWAS) were run for GDF15 (N=17,489) and NT-proBNP (N=16,963) levels in Generation Scotland. Protein measurements were transformed by rank-based inverse normalisation, regressed onto age, sex and 20 genetic principal components and scaled to have a mean of 0 and variance of 1. DNAm measurements in beta format were regressed onto age, sex and processing batch and scaled to have a mean of 0 and variance of 1. Houseman immune cell estimates were included as fixed effect covariates (42). Effect size estimates were obtained through Gibbs sampling over the posterior distribution, conditional on input data. The Gibbs protocol had 10,000 samples, with 5,000 samples of burn-in followed by a thinning of 5 samples to reduce autocorrelation. Methylation probes that had a posterior inclusion probability of ≥-95% were deemed to be significant for each protein.

### EpiScore development

Elastic net penalised regression was used to train EpiScores for GDF15 and Nt-pro-BNP levels. As Generation Scotland has extensive phenotyping and extant linkage to primary care and hospital records, EpiScores were first trained and tested in subsets of the full sample that were unrelated to one another to facilitate direct comparisons between EpiScore and protein levels in associations with incident diseases. EpiScores were then retrained on the full Generation Scotland sample and tested in LBC1936 – an external cohort. For both analyses, DNAm beta values were considered with missing CpG measurements mean imputed. To generate alternative versions of the EpiScores that can be applied to existing cohort studies with older Illumina array data (450k array), CpGs were filtered to the intersection of the 450k and EPIC array sites. A total of 760,838 EPIC array probes and 390,461 450k probes were available. CpG measurements were scaled to have a mean of 0 and variance of 1, prior to training. Protein measurements in training samples were transformed by rank-based inverse normalisation, regressed onto age, sex and 20 genetic principal components and scaled to have a mean of 0 and variance of 1. Penalised regression models were performed using Big Lasso (Version 1.5.1) in R (Version 4.0) (43). GDF15 and NT-proBNP protein levels were the outcomes. An elastic net penalty was specified (alpha=0.5). In the within-Generation Scotland analyses 10-fold cross validation was applied to select the lambda value that minimised the mean prediction error, whereas 20-fold cross validation was applied when training EpiScores in the full Generation Scotland sample.

A summary of the individuals with protein measurements available that were used to train and test EpiScores in the initial, within-Generation Scotland analyses is provided in **Supplementary Fig. 2**. Briefly, individuals that were part of the same family as disease cases in the test sample were excluded from the training sample. In the test subset of Generation Scotland, control individuals that were related to those in the training sample were excluded. A total of 8,207 individuals with GDF15 and 8,002 individuals with NT-proBNP measurements were therefore used to train EpiScores, while 2,954 individuals with GDF15 and 2,808 individuals with NT-proBNP measurements comprised the test samples. When retraining the EpiScores on the full Generation Scotland sample, there were 17,489 and 16,963 individuals available for the GDF15 and NT-proBNP EpiScores, respectively. **Supplementary Fig. 3** summarises the training and testing samples used, which included 500 individuals with NT-proBNP and 322 individuals with GDF15 measures in the external LBC1936 test set.

### EpiScore testing

To test EpiScores, the additional variance in protein levels that the EpiScores explained over a null model was quantified by running the following models:

Model 1: protein ∼ age + sex

Model 2: protein ∼ age + sex + protein EpiScore

The incremental variance (*R^2^*) in protein levels explained due to the protein EpiScore was calculated by subtracting the *R^2^* derived from model 1 from that in model 2. In these models, scaled, rank-based inverse normalised protein levels were used for testing. Pearson correlation coefficients were also calculated between GDF15 and NT-proBNP levels and their respective EpiScores in the test set and plotted. Protein EpiScores were tested using the described approach in both the Generation Scotland test subset (N≥2,808) and the individuals in Wave 4 of the LBC1936 external cohort that had measures of the proteins available (N_GDF_ = 322, N_NT-proBNP_ = 500). To assess the incremental variance that could be attributed to genetic architectures of the proteins, polygenic risk scores (PRS) for the proteins were calculated using genome-wide association study (GWAS) summary statistics generated in the Generation Scotland population via BOLT-LMM (44) **(see Supplementary Information)**. A summary of sentinel protein quantitative trait loci (pQTLs) identified by conditional and joint analyses (COJO) via Genome-wide Complex Trait Analysis (GCTA) software (45) for the GWAS results is available in **Supplementary Table 15**. PRS were derived using PRSice software (46). The PRS utilised pQTLs that had P < 5×10^−8^, with clumping (parameters: R^2^ = 0.25, distance = 250kb, p1 = 1). PRS were modelled in incremental variance assessments singularly and additively with the EpiScores in the test sets in relation to measured proteins.

### Cox proportional hazards analyses in Generation Scotland

Cox proportional hazards mixed effects regression models were used to assess the relationship between measured levels of GDF15 (N=17,489) and NT-proBNP (N=16,693) levels in the baseline Generation Scotland sample and four incident morbidities. The same model structure was also used in the test subset of the Generation Scotland sample where proteins and EpiScores were available for direct comparisons. All models were run using coxme (47) (Version 2.2-16) with a kinship matrix accounting for relatedness. Cases included those diagnosed after baseline who had died, in addition to those who received a diagnosis and remained alive. Controls were censored if disease free at time of death, or at the end of the follow-up period. Date of censoring was set to October 2020, which was the latest date of the GP data linkage information. Protein levels were rank-based inverse normalised and scaled to have a mean of 0 and variance of 1 prior to analyses. Basic models were run adjusting for age and sex. Fully-adjusted models further controlled for alcohol consumption (units consumed in the previous week); social deprivation (assessed by the Scottish Index of Multiple Deprivation (48)); body mass index (kilograms/height in metres squared); educational attainment (an 11-category ordinal variable) and a DNAm-based score for smoking status (49). Each of these covariates was sampled at baseline.

An FDR multiple testing correction P < 0.05 was applied to basic model associations across all diseases tested. Basic associations were considered to be significant if they had FDR P < 0.05. Associations in fully-adjusted models were considered to be significant if they had unadjusted P < 0.05. Proportional hazards assumptions were checked through Schoenfeld residuals (global test and a test for the protein variable) using the coxph and cox.zph functions from the survival package (50) (Version 3.2-7). For each association failing to meet the assumption (Schoenfeld residuals P < 0.05), a sensitivity analysis was run across yearly follow-up intervals. There were minimal differences in hazard ratios between follow-up periods that did not violate the assumption and those that did. All associations were therefore retained.

### COVID-19 analyses in Generation Scotland

Associations between measured levels of GDF15 and NT-proBNP and subsequent long-COVID (derived through CovidLife study survey 3 questionnaire (51)) or COVID-19 hospitalisation (derived through hospital linkage) were tested in the full Generation Scotland population. The preparation of the two binary outcome variables (long-COVID or hospitalisation due to COVID-19) is detailed in **Supplementary Information**. Logistic regression models with either hospitalisation (28 of 491 possible individuals) or long-COVID status (87 of 269 possible individuals) were run, with standardised (measured) proteins as the independent variables. Controls were defined as individuals that had COVID-19 but did not experience hospitalisation or long-COVID. Sex and age at COVID testing were adjusted for in the models. The latter was defined as the age at positive COVID-19 test or 1st January 2021 if COVID-19 test data were not available.

### EpiScore associations with brain health traits in LBC1936

As longitudinal cognitive testing and brain morphology measures are available in LBC1936, structural equation models (SEM) were used to examine the relationship between each EpiScore and brain health traits (cross-sectionally and longitudinally). Outcomes included: general cognitive ability (*g*), total brain volume, normal-appearing white matter volume, global grey matter volume and white matter hyperintensity volume. Cognitive test data were available at all measurement Waves (mean ages 70, 73, 76, 79 and 82) and brain magnetic resonance imaging (MRI) data were available from the second Wave (mean ages 73, 76, 79 and 82). Information on how the SEM analyses were constructed, with information on the number of individuals with cognitive and brain imaging measures at each Wave is included in **Supplementary Information**. Basic models were run with adjustment for age and sex, whereas fully-adjusted models included further covariates: DNAm-derived immune cell proportion estimates, DNAm-derived smoking score (49), self-reported alcohol consumption, BMI and the Scottish Index of Multiple Deprivation (52). Intercept (cross-sectional associations) and slope (longitudinal change) coefficients were extracted. A total of 1,091 individuals were modelled as part of the SEM analyses, with 895 individuals that had EpiScore measures available at Wave 1.

Individuals consented to share disease information from electronic health records from Wave 2 of the study onwards. Cox PH models were run to test associations between Wave 2 GDF15 and NT-proBNP EpiScores and incident dementia diagnoses after Wave 2 baseline, with basic adjustments for age and sex. The test population included 780 individuals who had dementia ascertainment and EpiScore information available at Wave 2, with 108 of these individuals having received a dementia diagnosis post-baseline (mean time-to-event 8.6 years [SD 3.4]). For the 108 incident cases, time-to-event was calculated using age at diagnosis. For controls who had died age at death was used for censoring, whereas age at the date of the dementia consensus meeting decision was taken forward for controls that remained alive.

## Data Availability

The source datasets from the cohorts that were analysed during the current study are not publicly available due to them containing information that could compromise participant consent and confidentiality. Data can be obtained from the data owners. Instructions for accessing Generation Scotland data can be found here: https://www.ed.ac.uk/generation-scotland/for-researchers/access; the GS Access Request Form can be downloaded from this site. Completed request forms must be sent to to be approved by the Generation Scotland Access Committee. According to the terms of consent for Generation Scotland participants access to data must be reviewed by the Generation Scotland Access Committee. Applications should be made to. LBC1936 data are available on request from the Lothian Birth Cohort Study University of Edinburgh https://www.ed.ac.uk/lothian-birth-cohorts/data-access-collaboration.
All correspondence and material requests should be sent to Riccardo Marioni at. All R code used in analyses is provided at: https://github.com/DanniGadd/EpiScores-GDF15-NT-proBNP. Latent Curve Growth Model (LCGM) LBC cognitive code that was adapted for these analyses is also available at: https://www.ed.ac.uk/lothian-birth-cohorts/summary-data-resources.

https://github.com/DanniGadd/EpiScores-GDF15-NT-proBNP

https://www.ed.ac.uk/lothian-birth-cohorts/summary-data-resources.

https://www.ed.ac.uk/generation-scotland/for-researchers/access

https://www.ed.ac.uk/lothian-birth-cohorts/data-access-collaboration

## List of abbreviations

BMI: Body mass index
CI: Confidence interval
COJO: Conditional and joint analyses
COVID-19: Coronavirus disease 2019
CpG: Cytosine-phosphate-guanine
DNAm: DNA methylation
EpiScore: Epigenetic score
EWAS: Epigenome-wide association study
FDR: False discovery rate
GCTA: Genome-wide Complex Trait Analysis
GDF15: Plasma growth differentiation factor 15
GWAS: Genome-wide association study
HbA1c: Glycated haemoglobin
HR: Hazard ratio
ICD: International Classification of Diseases codes
IHD: Ischaemic heart disease
IgG: Immunoglobulin G
MRI: Magnetic resonance imaging
NT-proBNP: N-terminal pro-B-type natriuretic peptide
LBC1936: The Lothian Birth Cohort of 1936
OR: odds ratio
PH: Proportional hazards
PIP: Posterior inclusion probability
pQTL: Protein quantitative trait loci
PRS: Polygenic risk score
SD: Standard deviation SE - Standard error
SEM: Structural equation model

## Declarations

### Ethics approval and consent to participate

All components of Generation Scotland received ethical approval from the NHS Tayside Committee on Medical Research Ethics (REC Reference Number: 05/S1401/89). Generation Scotland has also been granted Research Tissue Bank status by the East of Scotland Research Ethics Service (REC Reference Number: 20-ES-0021), providing generic ethical approval for a wide range of uses within medical research.

LBC1936 ethical approval was obtained from the Multicentre Research Ethics Committee for Scotland (Wave 1, MREC/01/0/56), the Lothian Research Ethics Committee (Wave 1, LREC/2003/2/29), and the Scotland A Research Ethics Committee (Waves 2-5, 07/MRE00/58). All participants provided written informed consent.

## Consent for publication

Not applicable.

## Availability of data and materials

The source datasets from the cohorts that were analysed during the current study are not publicly available due to them containing information that could compromise participant consent and confidentiality. Data can be obtained from the data owners. Instructions for accessing Generation Scotland data can be found here: https://www.ed.ac.uk/generation-scotland/for-researchers/access; the ‘GS Access Request Form’ can be downloaded from this site. Completed request forms must be sent to to be approved by the Generation Scotland Access Committee. According to the terms of consent for Generation Scotland participants, access to data must be reviewed by the Generation Scotland Access Committee. Applications should be made to.

LBC1936 data are available on request from the Lothian Birth Cohort Study, University of Edinburgh https://www.ed.ac.uk/lothian-birth-cohorts/data-access-collaboration.

All correspondence and material requests should be sent to Riccardo Marioni at. All R code used in analyses is provided at: https://github.com/DanniGadd/EpiScores-GDF15-NT-proBNP. Latent Curve Growth Model (LCGM) LBC cognitive code that was adapted for these analyses is also available at: https://www.ed.ac.uk/lothian-birth-cohorts/summary-data-resources.

## Authors’ contributions

D.A.G., and R.E.M. were responsible for the conception and design of the study. D.A.G. carried out all data analyses. D.A.G. and R.E.M. drafted the article. O.C., H.S., E.B., Y.C., R.F.H., D.M., C.F.R., and D.L.Mc. contributed to methodology. A.C., R.F., S.W.M., R.M.W., K.L.E., S.E.H., D.P., A.M.M., C.H., T.R., R.M.W., P.W. and N.S. contributed to the data collection and preparation. D.P., A.T., J.C. and S.R.C. collected and processed LBC1936 data. M.E.B., J.M.W., M.D.C.V.H., and S.M.M., derived brain imaging variables in LBC1936. R.E.M. supervised the project. All authors read and approved the manuscript.

## Acknowledgements

We acknowledge the participants of the Generation Scotland and Lothian Birth Cohort studies for their involvement in the data that have made this work possible. The authors thank all LBC1936 and Generation Scotland study research team members who have contributed, and continue to contribute, to ongoing studies.

