## Supplementary Figures for "DNAm scores for serum GDF15 and NT-proBNP levels associate with a range of traits affecting the body and brain"

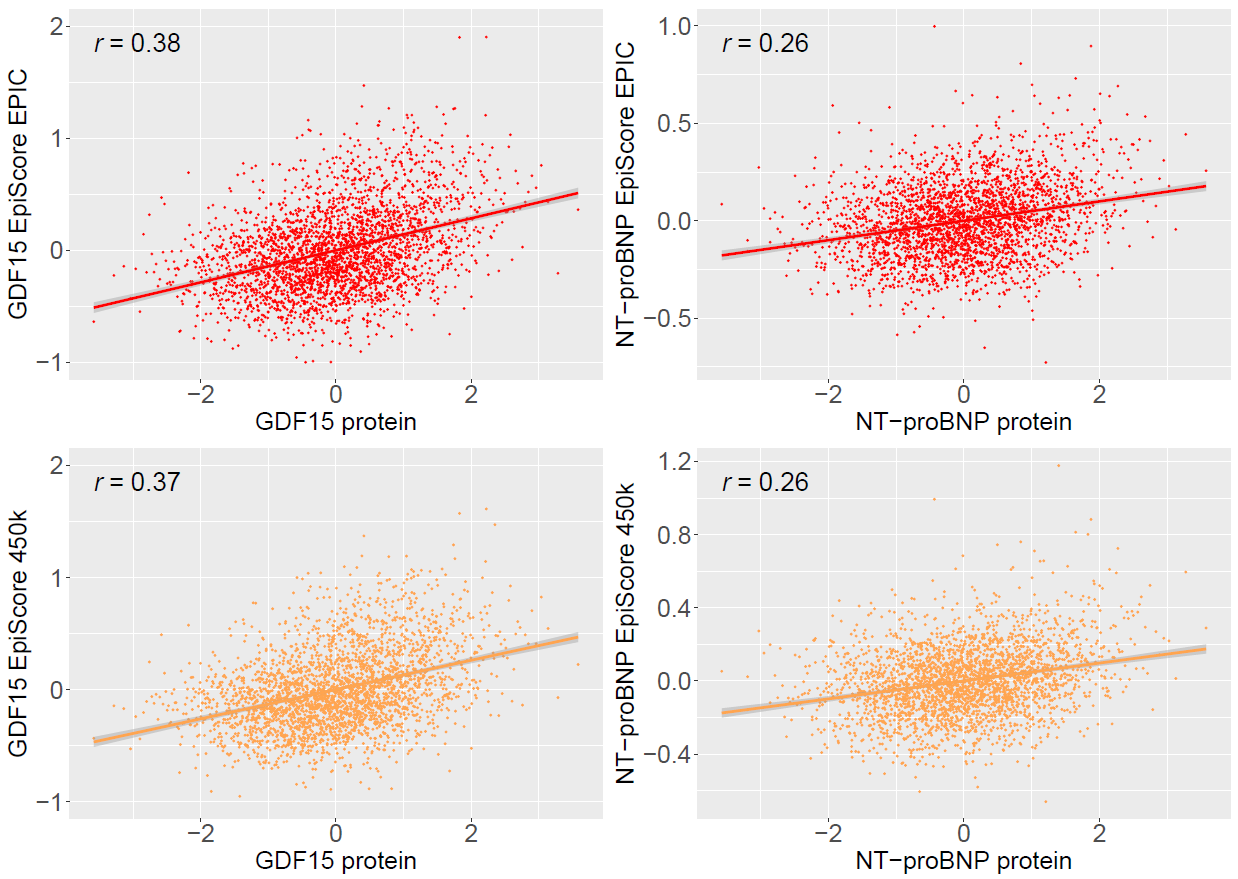

**Supplementary Figure 1. Pearson’s correlation structures between EpiScores and measured protein levels in Generation Scotland.** EpiScores for GDF15 and NT-proBNP trained using 760,838 possible CpGs from the EPIC array (red) and EpiScores trained using a subset of 390,461 CpGs available on the 450K array (orange) are presented. Test sample protein levels were rank-based inverse normalised and scaled to have a mean of 0 and standard deviation of 1. Pearson correlation coefficients are annotated in each case. The test sample included 2,954 individuals with GDF15 measurements and 2,808 individuals with Nt-proBNP measurements.

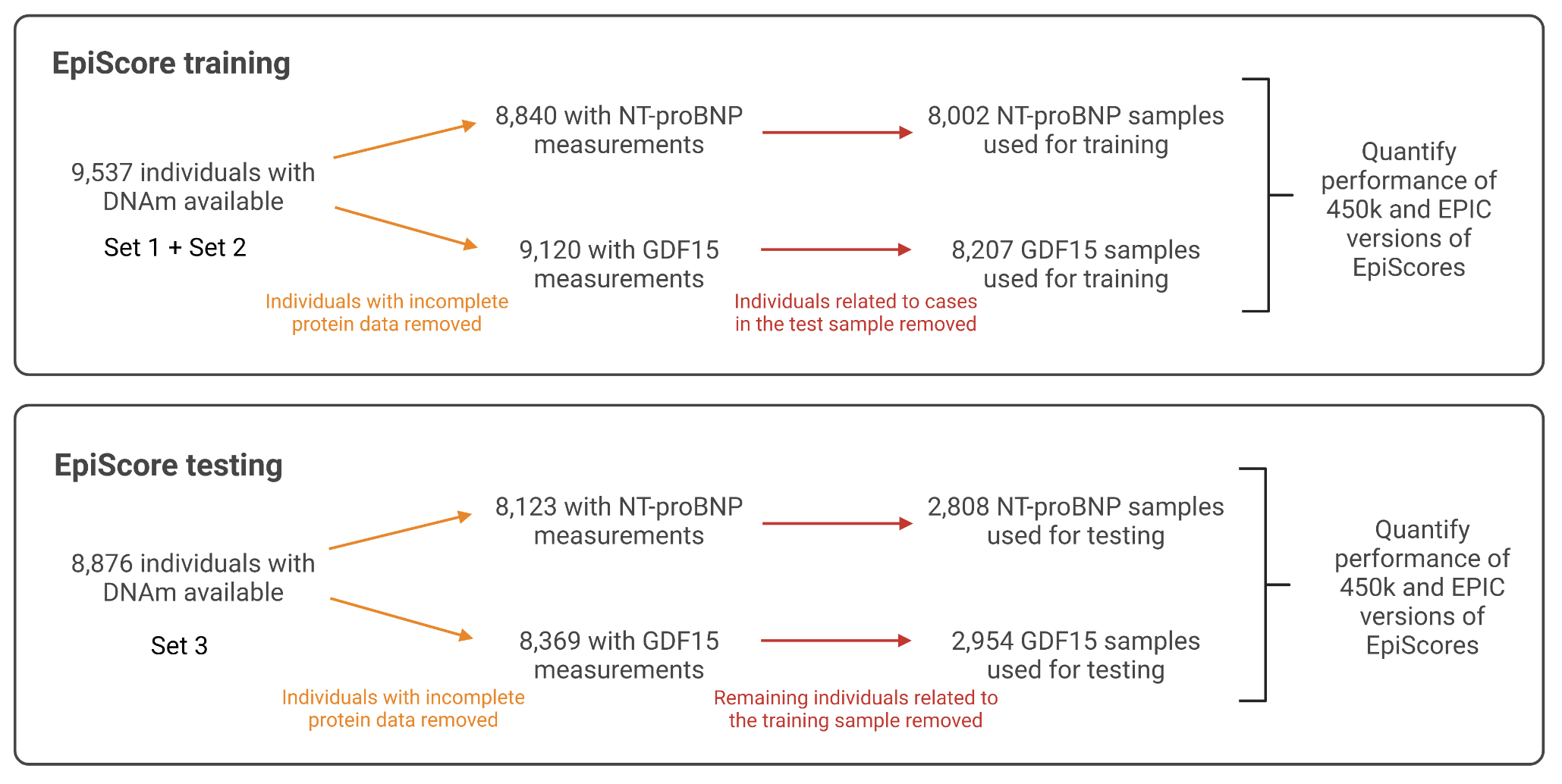

**Supplementary Figure 2. Initial training and testing sample selection for the generation of protein EpiScores for GDF15 and NT-proBNP in subsets of Generation Scotland.** DNA methylation from Sets 1 and 2 was available for training and Set 3 was available for testing. Individuals without complete protein measurements were removed in each case (orange). Samples were then subset to mitigate effects of relatedness (red). This involved first removing any individuals in the training sample that were related to cases in the test sample. The test sample was then subset to remove any individuals that were related to those in the training sample. This maximised cases available in the test sample for incident disease analyses, while also ensuring there were no individuals in the training set that were related to individuals in the testing set. Demographics for these populations are presented in **Supplementary Table 1**.

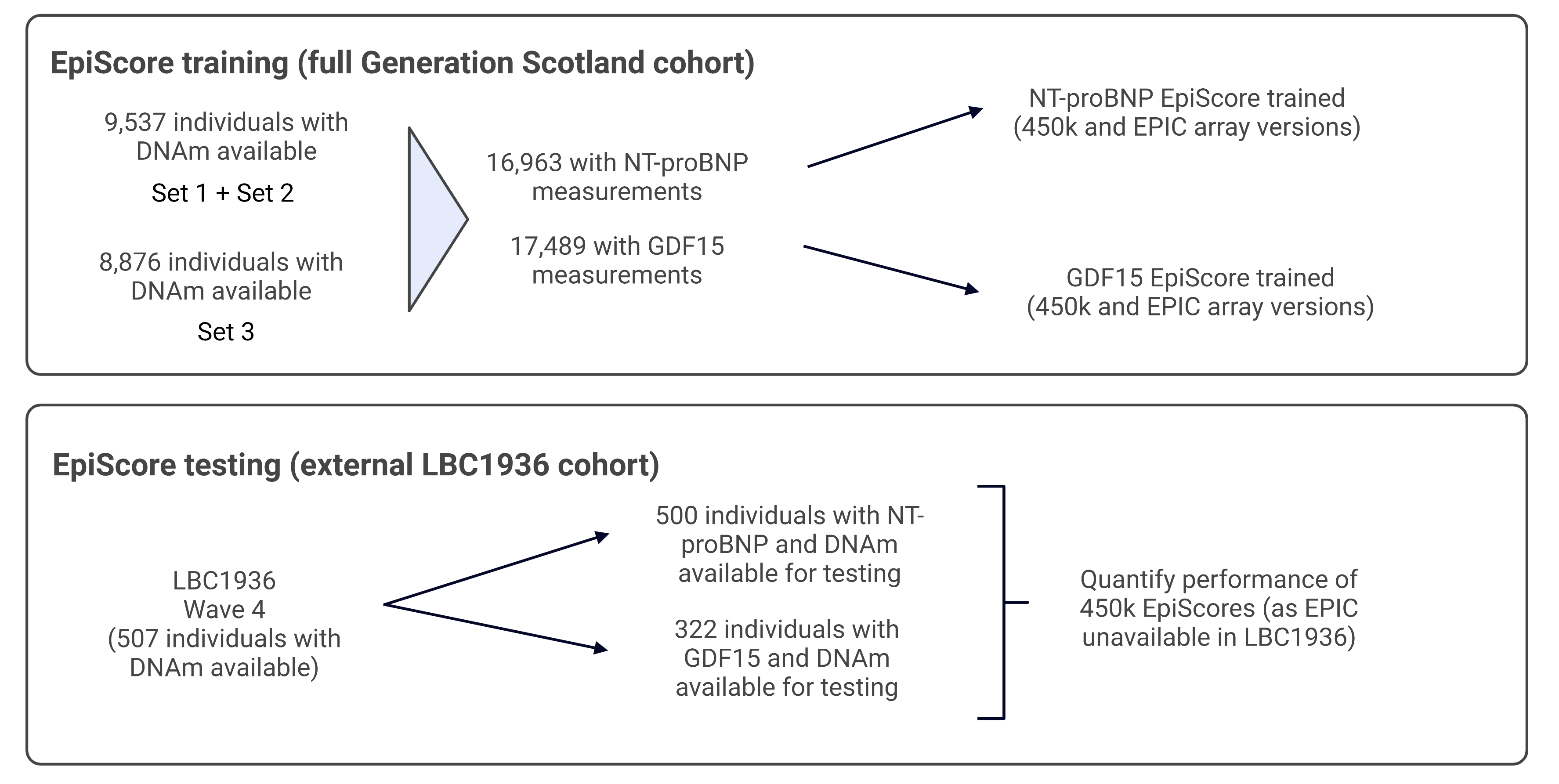

**Supplementary Figure 3. Sample selection for training protein EpiScores for GDF15 and NT-proBNP in the full Generation Scotland sample and testing in the external LBC1936 cohort.** DNA methylation from Sets 1, 2 and 3 were used for training in Generation Scotland. Measures of GDF15 and NT-proBNP at LBC1936 Wave 4 were available to test performance of the scores externally. Although versions of the EpiScores were trained using EPIC array and 450k sites, only the 450k was assessed in LBC1936 as measures of DNAm on the EPIC array were not available. Demographics for these populations are presented in **Supplementary Table 1**.
