## Supplementary Information for "DNAm scores for serum GDF15 and NT-proBNP levels associate with a range of traits affecting the body and brain"

**Quality control for DNA methylation in Generation Scotland**

DNA methylation data in Generation Scotland were generated in three separate sets, with 5,087 (Set 1), 4,450 (Set 2) and 8,877 (Set 3) individuals. Processing took place in 2017, 2019 and 2021, respectively. Set 1 and Set 3 included related individuals within each set. All individuals in Set 2 were unrelated to each other and to individuals in Set 1 (genetic relationship matrix (GRM) threshold <0.05). During DNA methylation quality control, CpG probes were filtered by removing those with low bead count (of <3) in ≥5% of samples or a high detection p-value (>0.05) in more than 5% of samples ^1,2^. Samples with a mismatch between predicted and recorded sex or ≥ 1% of CpGs with detection p-value > 0.05 were also removed in addition to saliva samples and genetic outliers ^3^. Cross-hybridising and single nucleotide polymorphism (SNP) associated probes and probes on the X and Y chromosomes were removed.

**GDF15 and NT-proBNP measurement**

Serum biomarker measurements were recorded at a single (second) thaw of stored aliquots. GDF-15 and NT-proBNP levels were measured on a cobas e411 analyser (Roche Diagnostics, Basel, Switzerland) using the manufacturer’s advised reagents and quality controls. The same assay was applied to Wave 4 serum samples in LBC1936 to generate protein measures.

**DNAm and protein measurements in the LBC1936 sample**

DNA methylation data were assessed through whole blood samples from LBC1936 with the Illumina HumanMethylation450 BeadChip (Illumina Inc., San Diego, CA, USA). Briefly, background correction was performed, followed by quality control. Probes with low quality (manually-identified), low detection rate (p > 0.01 for >5% of samples) or low call rate (p < 0.01 for <95% of probes) were excluded. Samples with a mismatch between predicted and recorded sex, or single nucleotide polymorphism controls probes were also removed.

**COVID-19 outcome preparation in Generation Scotland**

A binary variable for long-COVID based on self-reported COVID-19 duration from the CovidLife study survey 3 questionnaire (N=2,399 participating individuals in Generation Scotland) was used ^4^. Participants were asked about the total time they experienced symptoms in their first/only episode of illness, as well as the whole of their COVID-19 illness. The dataset is correct as of February 2021, when survey 3 was administered. Of 269 individuals that indicated they had been ill with COVID-19, 87 were classified as having long-COVID (> 4 weeks duration of symptoms after infection). Hospitalisation information, derived from the Scottish Morbidity Records (SMR01), was used to collate COVID-19 hospital admissions through ICD10 codes U07.1 (lab-confirmed COVID-19 diagnosis), and U07.2 (clinically-diagnosed COVID-19). This data linkage indicated that 491 individuals had COVID-19 diagnoses, with 28 of the individuals recorded as being hospitalised as a result of COVID-19. The mean years to follow-up was 11 years (SD 1 year) for both the long-COVID CovidLife survey and data linkage to hospitalisations due to COVID-19.

**PRS generation: GWAS sample preparation**

PRS for each protein were generated and modelled alongside the protein EpiScores to assess the additive variance in protein levels explained by epigenetic and genetic scores. To generate PRS for each protein, GWAS were run in the full Generation Scotland samples available (GDF15 = 17,489 individuals and NT-proBNP = 16,693 individuals). Genetic samples were genotyped using the Illumina HumanOmniExpressExome-8v1 chip and the Beadstudio-Gencall v3 genotype calling algorithm. Quality control excluded individuals with a call rate of ≤ 98% and SNPs with a call rate of ≤ 98%, HWE of ≤ 1x10-06 and MAF of ≤ 1%. Phasing of the genotyped SNPs was then carried out using SHAPEIT2 ^5^ and imputation was performed using the Haplotype Reference Consortium reference panel (HRC.r1-1) ^6^. After imputation, an additional quality control step excluded duplicate and monomorphic SNPs and SNPs with imputation quality scores < 0.4. After imputation and quality control, there were 24,161,581 SNPs available for analyses in GWAS.

**General cognitive ability measures in LBC1936**

Cognitive testing in the LBC1936 has been well-documented previously ^7–9^ and includes scores for 13 tests recorded across Waves 1-5 of the LBC1936 study. **Table 1** summarises the number of individuals with complete data for each of the measures of cognitive ability. Processing speed was measured through the inspection time, choice reaction time, digit symbol and symbol search tests. Memory capabilities were assessed through the verbal paired associates, logical memory and digit symbol backwards tests. The matrix reasoning, block design and spatial span tests evaluated visuospatial skills. Finally, verbal ability was recorded via the National and Wechsler adult reading tests and the verbal fluency test. Processing speed was measured using Digit-symbol Coding and Symbol Search (WAIS III-UK) ^10^ and two experimental tasks: Choice Reaction Time ^11^; and Inspection Time ^12^ Memory was measured using Verbal Paired Associates and Logical Memory (WMSIII-UK) ^10^ and Digit-span Backwards (WAIS-IIIUK). Visuospatial ability was measured using Block Design and Matrix Reasoning (WAIS-IIIUK) and Spatial Span (Forwards and Backward) (WMS-IIIUK). Verbal ability was measured using the National Adult Reading Test (NART) ^13^ the Wechsler Test of Adult Reading (WTAR) ^14^ and Verbal Fluency ^15^.

**MRI measures in LBC**

Magnetic resonance imaging (MRI) data collection and processing in the LBC1936 has been documented previously ^16^. Briefly, the same 1.5T GE Sigma Horizon HDx clinical scanner was utilised to measure T1-weighted (voxel sizes: 1x1x1.3mm) and T2-weighted (voxel sizes: 1x1x2mm) and fluid-attenuated inversion recovery-weighted images (voxel sizes: 1x1x4mm) at each time point ^16^. The structural MRI measures included as indictors of brain health in the present study included: total brain volume, normal appearing white matter volume, grey matter volume and white matter hyperintensity volume. Intracranial volumes were determined semi-automatically using Analyze 11.0TM and included as a covariate in all brain imaging analyses. Total brain and normal appearing white matter volumes were calculated via a semi-automated multi-spectral fusion approach ^17^. The availability of MRI measures used in the study at each Wave are summarised in **Table 2**. These measures were recorded across Waves 2-5 of the LBC1936, representing 9.5 years in total.

| Test | Wave | N | Mean | SD | Missing | Range |
| --- | --- | --- | --- | --- | --- | --- |
| Digit symbol backwards | 1 | 1090 | 7.734862 | 2.262883 | 1 | 2 - 14 |
| Digit symbol backwards | 2 | 866 | 7.814088 | 2.289223 | 225 | 2 - 14 |
| Digit symbol backwards | 3 | 695 | 7.768345 | 2.373516 | 396 | 2 - 14 |
| Digit symbol backwards | 4 | 548 | 7.558394 | 2.177302 | 543 | 2 - 14 |
| Digit symbol backwards | 5 | 426 | 7.190141 | 2.33016 | 665 | 0 - 13 |
| Matrix Reasoning | 1 | 1086 | 13.49448 | 5.131404 | 5 | 4 - 24 |
| Matrix Reasoning | 2 | 863 | 13.17497 | 4.963975 | 228 | 3 - 25 |
| Matrix Reasoning | 3 | 689 | 13.03919 | 4.909048 | 402 | 3 - 25 |
| Matrix Reasoning | 4 | 535 | 12.90467 | 5.032877 | 556 | 1 - 25 |
| Matrix Reasoning | 5 | 418 | 12.92584 | 5.219568 | 673 | 3 - 25 |
| Block design | 1 | 1085 | 33.78894 | 10.3217 | 6 | 10 - 65 |
| Block design | 2 | 864 | 33.63773 | 10.07766 | 227 | 10 - 66 |
| Block design | 3 | 691 | 32.18379 | 9.951889 | 400 | 3 - 66 |
| Block design | 4 | 535 | 31.2 | 9.633898 | 556 | 0 - 63 |
| Block design | 5 | 420 | 29.9 | 9.599349 | 671 | 2 - 62 |
| Digit symbol | 1 | 1086 | 56.59945 | 12.93268 | 5 | 14 - 98 |
| Digit symbol | 2 | 862 | 56.40023 | 12.30992 | 229 | 15 - 94 |
| Digit symbol | 3 | 685 | 53.80876 | 12.93291 | 406 | 14 - 89 |
| Digit symbol | 4 | 535 | 51.24112 | 13.00777 | 556 | 12 - 89 |
| Digit symbol | 5 | 418 | 50.98086 | 12.78974 | 673 | 15 - 81 |
| Symbol search | 1 | 1084 | 24.75554 | 6.285176 | 7 | 2 - 49 |
| Symbol search | 2 | 862 | 24.60905 | 6.175262 | 229 | 3 - 45 |
| Symbol search | 3 | 687 | 24.60408 | 6.456211 | 404 | 3 - 53 |
| Symbol search | 4 | 528 | 22.73106 | 6.627499 | 563 | 2 - 50 |
| Symbol search | 5 | 415 | 22.21205 | 6.929831 | 676 | -1 - 40 |
| National adult reading test | 1 | 1089 | 34.48209 | 8.154522 | 2 | 1 - 50 |
| National adult reading test | 2 | 864 | 34.37963 | 8.175471 | 227 | 1 - 50 |
| National adult reading test | 3 | 695 | 35.02446 | 8.027024 | 396 | 5 - 49 |
| National adult reading test | 4 | 546 | 35.59158 | 8.192178 | 545 | 6 - 50 |
| National adult reading test | 5 | 426 | 36.05164 | 7.813241 | 665 | 8 - 49 |
| Logical memory | 1 | 1087 | 71.4609 | 17.96493 | 4 | 0 - 117 |
| Logical memory | 2 | 864 | 74.30208 | 17.87896 | 227 | 5 - 116 |
| Logical memory | 3 | 688 | 74.57703 | 19.20183 | 403 | 5 - 116 |
| Logical memory | 4 | 542 | 72.71033 | 20.39497 | 549 | 0 - 122 |
| Logical memory | 5 | 421 | 72.14727 | 21.52102 | 670 | 2 - 118 |
| Verbal paired associates | 1 | 1050 | 26.44286 | 9.132593 | 41 | 0 - 40 |
| Verbal paired associates | 2 | 843 | 27.18268 | 9.460875 | 248 | 0 - 40 |
| Verbal paired associates | 3 | 663 | 26.41026 | 9.557617 | 428 | 1 - 40 |
| Verbal paired associates | 4 | 497 | 27.14085 | 9.554612 | 594 | 0 - 40 |
| Verbal paired associates | 5 | 380 | 27.36842 | 9.542077 | 711 | 0 - 40 |
| Inspection time | 1 | 1041 | 112.1402 | 10.99998 | 50 | 46 - 140 |
| Inspection time | 2 | 838 | 111.2232 | 11.78852 | 253 | 32 - 140 |
| Inspection time | 3 | 654 | 110.1361 | 12.55365 | 437 | 49 - 136 |
| Inspection time | 4 | 465 | 106.9591 | 13.60133 | 626 | 37 - 132 |
| Inspection time | 5 | 382 | 106.034 | 12.72447 | 709 | 61 - 136 |
| Four choice reaction time | 1 | 1084 | 0.642107 | 0.08578 | 7 | 0.45 - 1.13 |
| Four choice reaction time | 2 | 865 | 0.649272 | 0.089842 | 226 | 0.45 - 1.29 |
| Four choice reaction time | 3 | 685 | 0.678844 | 0.102754 | 406 | 0.46 - 1.31 |
| Four choice reaction time | 4 | 543 | 0.706031 | 0.113613 | 548 | 0.46 - 1.34 |
| Four choice reaction time | 5 | 423 | 0.722026 | 0.120251 | 668 | 0.49 - 1.24 |
| Spatial span | 1 | 1084 | 14.71771 | 2.831954 | 7 | 5 - 24 |
| Spatial span | 2 | 861 | 14.69454 | 2.762006 | 230 | 5 - 23 |
| Spatial span | 3 | 690 | 14.61594 | 2.728452 | 401 | 6 - 22 |
| Spatial span | 4 | 536 | 14.1306 | 2.718183 | 555 | 7 - 23 |
| Spatial span | 5 | 421 | 13.89311 | 2.851144 | 670 | 3 - 21 |
| Verbal fluency | 1 | 1087 | 42.41766 | 12.53791 | 4 | 10 - 83 |
| Verbal fluency | 2 | 865 | 43.18035 | 12.94021 | 226 | 6 - 90 |
| Verbal fluency | 3 | 696 | 42.89799 | 12.7646 | 395 | 6 - 86 |
| Verbal fluency | 4 | 547 | 43.6106 | 13.3337 | 544 | 13 - 83 |
| Verbal fluency | 5 | 426 | 43.55399 | 12.68971 | 665 | 4 - 80 |
| Wechsler test of reading | 1 | 1089 | 41.02112 | 7.174846 | 2 | 8 - 50 |
| Wechsler test of reading | 2 | 864 | 41.01042 | 6.966806 | 227 | 5 - 50 |
| Wechsler test of reading | 3 | 694 | 41.09222 | 7.021724 | 397 | 8 - 50 |
| Wechsler test of reading | 4 | 546 | 41.62821 | 7.033279 | 545 | 12 - 50 |
| Wechsler test of reading | 5 | 426 | 42.19014 | 6.606521 | 665 | 18 - 50 |

**Table 1. Summary of cognitive ability components in the LBC1936 cohort.** Each cognitive score components used in the derivation of the cognitive ability phenotype modelled in SEM is summarised for LBC1936.

| Measure | Wave | N | Mean | SD | Missing | Range |
| --- | --- | --- | --- | --- | --- | --- |
| Intrcranial volume (cm^3^) | 2 | 697 | 1450635 | 140461.6 | 394 | 1059966 - 1876420 |
| Total brain volume (cm^3^) | 2 | 658 | 988971.4 | 89437.39 | 433 | 730751.43 - 1264608.52 |
| Total brain volume (cm^3^) | 3 | 476 | 973752.1 | 90629.47 | 615 | 712210.43 - 1248294.06 |
| Total brain volume (cm^3^) | 4 | 382 | 963528.4 | 89079.45 | 709 | 702959.53 - 1227385.39 |
| Total brain volume (cm^3^) | 5 | 301 | 947223.7 | 87099.39 | 790 | 702114.86 - 1182018.72 |
| White matter hyperintensity volume (cm^3^) | 2 | 672 | 12062.54 | 12840.57 | 419 | 0 - 98378 |
| White matter hyperintensity volume (cm^3^) | 3 | 476 | 16340.38 | 15621.06 | 615 | 36 - 92080 |
| White matter hyperintensity volume (cm^3^) | 4 | 382 | 19779.09 | 17285.41 | 709 | 552 - 101238 |
| White matter hyperintensity volume (cm^3^) | 5 | 301 | 21458.3 | 18246.36 | 790 | 786 - 108132 |
| Grey matter volume (cm^3^) | 2 | 658 | 471555.4 | 44707.8 | 433 | 366423.96 - 616236.61 |
| Grey matter volume (cm^3^) | 3 | 476 | 464678.1 | 43462.64 | 615 | 347811.05 - 593406.84 |
| Grey matter volume (cm^3^) | 4 | 382 | 463070.4 | 45022.25 | 709 | 327650.85 - 624006.15 |
| Grey matter volume (cm^3^) | 5 | 301 | 445530.7 | 43347.71 | 790 | 273419.98 - 580192.77 |
| Normal appearing white matter volume (cm^3^) | 2 | 658 | 475009.9 | 50663.64 | 433 | 301681.36 - 668774.71 |
| Normal appearing white matter volume (cm^3^) | 3 | 476 | 463466.3 | 53436.68 | 615 | 279666.83 - 651542.15 |
| Normal appearing white matter volume (cm^3^) | 4 | 382 | 455876.9 | 54000 | 709 | 222377.18 - 668132.84 |
| Normal appearing white matter volume (cm^3^) | 5 | 301 | 453124 | 54354.07 | 790 | 256467.84 - 638024.94 |

**Table 2. Summary of MRI measures in the LBC1936 cohort.** Each MRI phenotype modelled in SEM is summarised for LBC1936. Brain imaging data are available from Wave 2 onwards.

**Structural equation modelling in LBC1936**

The Lavaan R package (Version 0.6-12) ^18^ was used to generate latent variables through growth curve modelling that were included in the structural equation modelling (SEM) framework. Intercepts and slopes for each of the 13 cognitive tests were generated using cognitive testing data from 5 waves of testing. Loadings were fixed to 1 for intercepts and to mean lag time (0, 2.98, 6.75, 9.82, and 12.54) between waves for slopes. A first-order hierarchical model of general cognitive ability (intercept) and change (slope) using latent growth curve modelling with factor of curves specification ^19^. This was done in accordance with previous extraction of cognitive ability based on the correlational structure of cognitive domains in this cohort population ^20,21^. Residual covariances between tests in the same cognitive domain were included in the model ^22^. Negative residual variances were fixed to zero and scaling was performed according to the first variable (marker method). Growth curve models in a SEM framework were also used to produce latent variables baseline (intercept) and change (slope) for each MRI measure. Measures of each MRI measure were loaded onto a latent variable for intercept with loadings set to 1 for intercept for each wave. The measurements for each brain trait were loaded onto a latent variable for slope with each loading fixed at the mean lag time between the corresponding wave and baseline. The loadings for cognitive and brain imaging traits are described in **Tables 3-4**. Models employed full information maximum likelihood to include all data available. Model fit measures include Tucker-Lewis index (TLI), confirmatory factor index (CFI), root mean squared error approximation (RMSEA) and standardised root mean squared residual (SRMR). The thresholds for each of these measures that were considered acceptable were: > 0.95 for CFI and TLI, < 0.08 for SRMR and < 0.06 for RMSEA. Fit measures for model measurements of general cognition in LBC1936 were CFI = 0.958, TLI = 0.957, SRMR = 0.061 and RMSEA = 0.029. Equivalent fit measures for MRI model measurements are included in **Table 5**.

| Cohort | Test | Intercept | Slope |
| --- | --- | --- | --- |
| LBC1936 | Block design | 0.702 | 0.975 |
| LBC1936 | Matrix reasoning | 0.793 | 1 |
| LBC1936 | Span total | 0.642 | 1 |
| LBC1936 | NART | 0.685 | 0.474 |
| LBC1936 | WTAR | 0.679 | 0.438 |
| LBC1936 | Verbal fluency | 0.534 | 0.825 |
| LBC1936 | Verbal paired associates | 0.543 | 0.689 |
| LBC1936 | Logical memory | 0.615 | 0.764 |
| LBC1936 | Digit backwards | 0.722 | 1 |
| LBC1936 | Symbol search | 0.788 | 0.94 |
| LBC1936 | Digit symbol | 0.668 | 0.927 |
| LBC1936 | Inspection time | 0.491 | 0.92 |
| LBC1936 | Four choice reaction time | 0.526 | 0.889 |

**Table 3. Loadings for cognitive test contributions to the generation of cognitive ability across LBC1936 individuals used in the SEM framework.**

| Measure | Wave | Intecept | Slope |
| --- | --- | --- | --- |
| Grey matter volume | 2 | 0.977 | 0.000 |
| Grey matter volume | 3 | 0.998 | 0.140 |
| Grey matter volume | 4 | 0.988 | 0.252 |
| Grey matter volume | 5 | 0.980 | 0.350 |
| Total Brian volume | 2 | 0.988 | 0.000 |
| Total Brian volume | 3 | 1.003 | 0.076 |
| Total Brian volume | 4 | 1.007 | 0.139 |
| Total Brian volume | 5 | 1.018 | 0.197 |
| Normal appearing white matter volume | 2 | 0.988 | 0.000 |
| Normal appearing white matter volume | 3 | 0.940 | 0.198 |
| Normal appearing white matter volume | 4 | 0.880 | 0.337 |
| Normal appearing white matter volume | 5 | 0.834 | 0.447 |
| White matter hyperintensity volume | 2 | 0.631 | 0.000 |
| White matter hyperintensity volume | 3 | 1.077 | 0.185 |
| White matter hyperintensity volume | 4 | 1.137 | 0.355 |
| White matter hyperintensity volume | 5 | 1.175 | 0.513 |

**Table 4. Loadings for MRI measures at each Wave of LBC1936 used in the SEM framework.**

| Measure | CFI | TLI | RMSEA | SRMR |
| --- | --- | --- | --- | --- |
| Grey matter volume | 0.941 | 0.924 | 0.101 | 0.035 |
| Total Brain volume | 0.992 | 0.990 | 0.044 | 0.016 |
| Normal appearing white matter volume | 0.994 | 0.992 | 0.034 | 0.017 |
| White matter hyperintensity volume | 0.937 | 0.919 | 0.099 | 0.062 |

**Table 5. Fit for model measurements for MRI measures used in SEM framework.**
